# Seroprevalence and Correlates of SARS-CoV-2 Antibodies in Healthcare Workers in Chicago

**DOI:** 10.1101/2020.09.11.20192385

**Authors:** John T. Wilkins, Elizabeth L. Gray, Amisha Wallia, Lisa R. Hirschhorn, Teresa R. Zembower, Joyce Ho, Naomi Kalume, Ojoma Agbo, Alex Zhu, Laura J. Rasmussen-Torvik, Sadiya S. Khan, Mercedes Carnethon, Mark Huffman, Charlesnika T. Evans

## Abstract

**Background:** Identifying factors associated with SARS-CoV-2 infection among healthcare workers (HCW)s may help health systems optimize SARS-CoV-2 infection control strategies. Methods: We conducted a cross-sectional analysis of baseline data from the Northwestern HCW SARS-CoV-2 Serology Cohort Study. The Abbott Architect Nucleocapsid IgG assay was used to determine seropositivity. Logistic regression models (unadjusted and adjusted for demographics and self-reported community exposure to COVID-19) were fit to quantify the associations between occupation group, healthcare delivery tasks, and community exposure and seropositive status.

**Results:** 6,510 HCWs, including 1,794 nurses, and 904 non-patient facing administrators participated. The majority were women (79.6%), 74.9% were white, 9.7% were Asian, 7.3% were Hispanic and 3.1% were Black. The crude prevalence rate of seropositivity was 4.8% (95% confidence interval (CI): 4.6%-5.2%). Out-of-hospital exposure to COVID-19 occurred in 9.3% of HCWs and was strongly associated with seropositivity (OR=4.7, 95% CI: 3.5-6.4). When compared to administrators, nursing was the only occupation group with a significantly higher adjusted-odds (OR: 1.9, 95% CI: 1.3-2.9) of seropositivity. Exposure to COVID-19 patients was reported by 37.8% of participants and was associated with higher positivity than those not exposed (OR= 2.2, 95% CI: 1.6-3.0). Being exposed to patients receiving high-flow oxygen therapy, and hemodialysis also remained significantly associated with a 45% and 57% higher odds for seropositive status, respectively.

**Conclusions:** Exposure to COVID-19 patients, and longer duration patient therapies were each associated with higher risk for seropositive status; however, the community burden of COVID-19 remains a significant source of exposure to SARS CoV-2 infection among HCWs in Chicago.

## Introduction

Healthcare workers (HCWs) have provided essential front-line care for patients throughout the COVID-19 pandemic at considerable personal risk. Data from the Centers for Disease Control and Prevention (CDC) found that 11% of the total number of reported COVID-19 cases in the US were HCWs.(1) Supply chain problems that limited the availability of personal protective equipment (PPE) early in the pandemic have stabilized, but community transmission across the US has continued to rise.(2) To protect the essential workforce that delivers care, it remains a high priority to identify factors associated with SARS-CoV-2 infection in healthcare settings.

The city of Chicago experienced an early, prolonged surge in COVID-19 cases and was second only to the Northeastern tri-state region in cases, hospitalizations, and deaths through the end of June 2020.(3) Despite the significant burden of disease, aggressive public health mitigation efforts and system-wide inpatient expansion efforts ensured that hospital bed capacity was not exceeded and PPE supplies were not exhausted. Thus, Chicago-area health systems may serve as models for what risks are likely to be observed moving forward assuming continued public health mitigation efforts, bed expansion efforts, and adequate PPE supply lines.

We established the Northwestern Healthcare Worker SARS-CoV-2 Serology Study Cohort in May 2020 to determine the prevalence and correlates of anti-SARS-CoV-2 IgG status. The objective of our study was to describe the prevalence of SARS-CoV-2 seropositivity and correlates by occupational categories, clinical tasks, and sociodemographic characteristics. We collected information about community and household exposures to describe the relative contribution of out-of-hospital SARS-CoV-2 exposures to seropositivity among HCWs. In this manuscript, we report the cross-sectional baseline findings from the cohort study. We hypothesized that HCWs that participated in aerosolizing procedures, those with high COVID-19 patient exposure, and self-reported out-of-hospital exposures would have higher prevalence rates of anti-SARS-CoV-2 antibodies than those without these exposures.

## Methods

### Study Design and Setting

This investigation is part of an ongoing, prospective cohort study of SARS-CoV-2 in patient-facing and non-patient facing HCWs from a large, tertiary academic healthcare system that included 10 hospitals, 18 immediate care centers, and 325 outpatient practices in the Chicago area and surrounding IL suburbs. The largest hospital in the health system is located in downtown Chicago, whereas the other 9 regional centers are in the west, northwest, and north suburbs of Chicago. Affiliated outpatient practices and immediate care centers are located in downtown Chicago and the near suburbs.

At the time the employer (Northwestern Medicine (NM)) was planning to initiate an employer-sponsored benefit of free SARS-CoV-2 serology assessment, our team approached them about performing a research study. All institutional HCWs were eligible for participation in the benefit (see Table 1 and Supplemental Table 1). Participation in the research study was not required to receive serology testing results. This study was approved by the Northwestern University Institutional Review Board and all participants gave written informed consent. All HCWs (employees and physician members of affiliated outpatient practices) were invited to join the study May 28, 2020-June 30, 2020. Outreach consisted of existing methods of health care system communication including emails and information banners imbedded in the health system clinical information website. The email invitation specified 41 locations across Chicago and suburban areas where HCWs could obtain serological testing for SARS-CoV-2 and included information about the cohort study and an electronic link to consent and enroll. Testing was available through July 8, 2020. Due to low enrollment from environmental services, food service, and patient transportation groups, research team members (JTW, CTE) conducted one in-person recruitment briefing with this group. Twenty-one individuals in these occupation groups subsequently enrolled in the study.

**Table 1:** Characteristics of Participants by Occupation Groups

| Characteristics |  | Overall | RN | MD | Administrative Role | Other Occupations<sup>a</sup> |
| --- | --- | --- | --- | --- | --- | --- |
| **n** |  | 6510 | 1794 | 1260 | 904 | 2552 |
| **Age Category (%)** | 18-29 | 1304 (20.0) | 557 (31.0) | 128 (10.2) | 84 ( 9.3) | 535 (21.0) |
|  | 30-39 | 2208 (33.9) | 519 (28.9) | 527 (41.8) | 296 (32.7) | 866 (33.9) |
|  | 40-49 | 1368 (21.0) | 326 (18.2) | 310 (24.6) | 197 (21.8) | 535 (21.0) |
|  | 50-59 | 1042 (16.0) | 245 (13.7) | 177 (14.0) | 225 (24.9) | 395 (15.5) |
|  | 60+ | 588 ( 9.0) | 147 ( 8.2) | 118 ( 9.4) | 102 (11.3) | 221 ( 8.7) |
| **Gender (%)** | Female<sup>b</sup> | 5180 (79.6) | 1701 (94.8) | 682 (54.1) | 699 (77.3) | 2098 (82.2) |
|  | Male | 1330 (20.4) | 93 ( 5.2) | 578 (45.9) | 205 (22.7) | 454 (17.8) |
| **Race (%)** | Asian | 634 ( 9.7) | 125 ( 7.0) | 283 (22.5) | 50 ( 5.5) | 176 ( 6.9) |
|  | Hispanic/Latino | 477 ( 7.3) | 105 ( 5.9) | 49 ( 3.9) | 63 ( 7.0) | 260 (10.2) |
|  | Multiracial | 136 ( 2.1) | 23 ( 1.3) | 33 ( 2.6) | 20 ( 2.2) | 60 ( 2.4) |
|  | Non-hispanic Black | 201 ( 3.1) | 22 ( 1.2) | 25 ( 2.0) | 36 ( 4.0) | 118 ( 4.6) |
|  | Non-hispanic White | 4877 (74.9) | 1496 (83.4) | 843 (66.9) | 717 (79.3) | 1821 (71.4) |
|  | Other/NA<sup>c</sup> | 185 ( 2.8) | 23 ( 1.3) | 27 ( 2.1) | 18 ( 2.0) | 117 ( 4.6) |
| **Diabetes (%)** | Yes | 191 ( 2.9) | 49 ( 2.7) | 22 ( 1.7) | 28 ( 3.1) | 92 ( 3.6) |
|  | No | 6189 (95.1) | 1731 (96.5) | 1229 (97.5) | 868 (96.0) | 2361 (92.5) |
|  | NA | 130 ( 2.0) | 14 ( 0.8) | 9 ( 0.7) | 8 ( 0.9) | 99 ( 3.9) |
| **Obesity (%)** | Yes | 982 (15.1) | 283 (15.8) | 89 ( 7.1) | 171 (18.9) | 439 (17.2) |
|  | No | 5382 (82.7) | 1492 (83.2) | 1163 (92.3) | 725 (80.2) | 2002 (78.4) |
|  | NA | 146 ( 2.2) | 19 ( 1.1) | 8 ( 0.6) | 8 ( 0.9) | 111 ( 4.3) |
| **High Blood Pressure (%)** | Yes | 800 (12.3) | 197 (11.0) | 122 ( 9.7) | 151 (16.7) | 330 (12.9) |
|  | No | 5581 (85.7) | 1575 (87.8) | 1130 (89.7) | 749 (82.9) | 2127 (83.3) |
|  | NA | 129 ( 2.0) | 22 ( 1.2) | 8 ( 0.6) | 4 ( 0.4) | 95 ( 3.7) |

<sup>a</sup>For complete list of occupational groups included in the “other occupations” group please see Supplemental Table 1.
<sup>b</sup>Includes <10 that did not self-identify.
<sup>c</sup>NA: Not answered

A total of 38,127 NM HCWs received email invitations to participate in the employee benefit to have serology checked: 18,985 (49.8%) participated in the employer-sponsored serology benefit. Among the latter group, 6,714 (35.4%) enrolled in the cohort study. After exclusions for withdrawal of consent, no baseline survey data completed, or inability of the research team to verify the identity of the participant or view serology results (n=204), 6,510 participants comprised the final study sample. (Supplemental Figure 1)

### SARS-CoV-2 IgG Assay Testing

Blood samples were collected by a trained phlebotomist. The SARS-CoV-2 IgG assay on the high-throughput ARCHITECT i2000SR Immunoassay System from Abbott Laboratories (Abbott Park, IL) was used. The SARS-CoV-2 IgG assay is a qualitative, chemiluminescent microparticle immunoassay that identifies whether human serum or plasma have IgG antibodies to SARS-CoV-2 nucleocapsid antigen. Performance characteristics for this assay are reported to be 100% positive agreement at ≥ 14 days post-symptom onset in those with confirmed COVID-19 and 99.6% negative agreement in those without COVID-19.(4)

### Health System Infection Control Procedures

Since January 2020, droplet isolation precautions were used on all patients at NM with known or suspected COVID-19. N95 respirators were recommended for aerosol-generating procedures. Universal masking was initiated in late March. NM had adequate PPE available for use by all staff at all times. In early March, COVID-19 inpatients were cared for in COVID-19 floors and ICUs. Remote working was mandated whenever possible for all HCWs. NM inpatient COVID-19 census and Chicago cases are shown in Figure 1.

**Figure 1:**
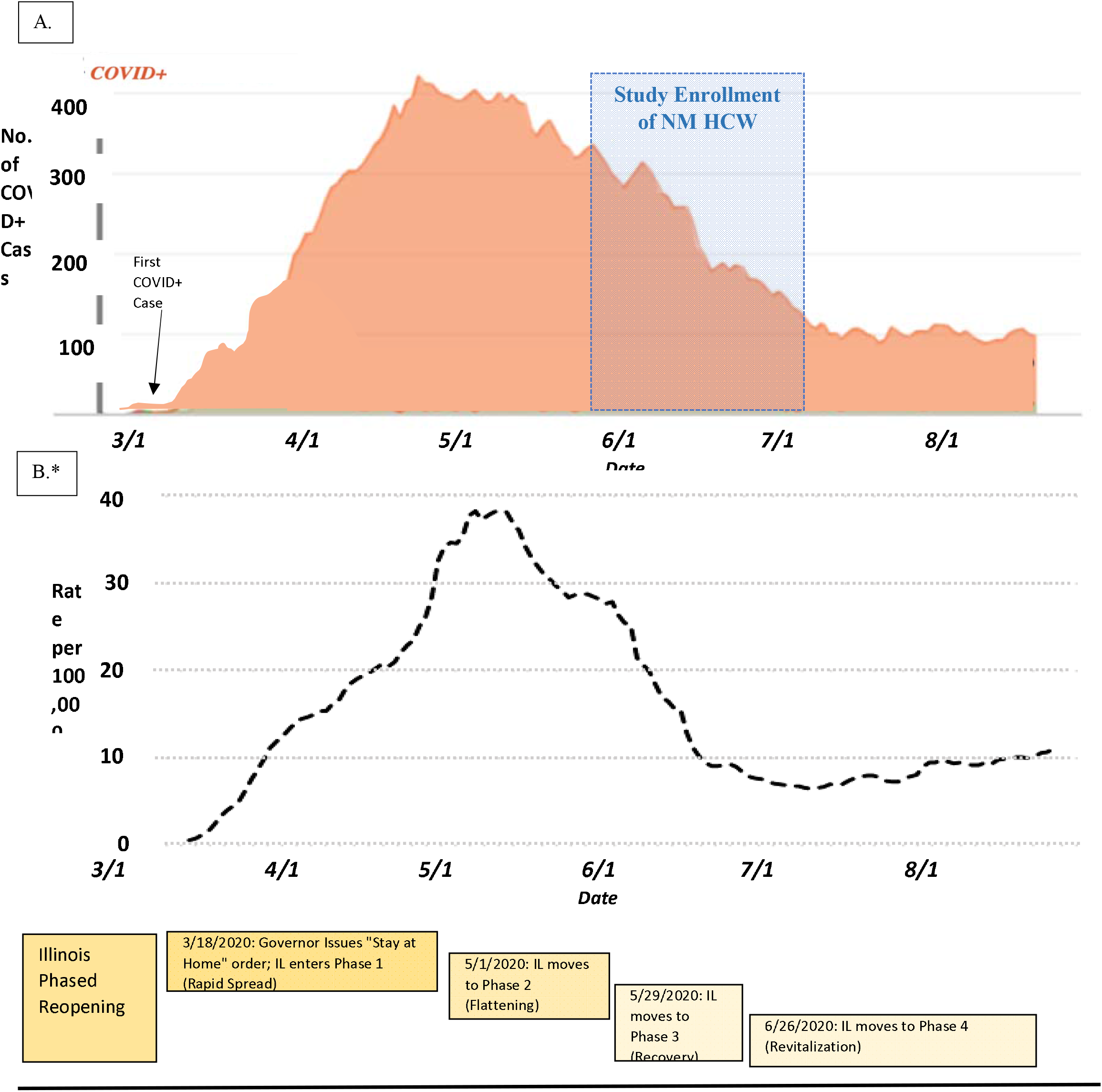
Timeline of Northwestern Medicine COVID-19 Inpatient Census, Chicago Case Rate, and state government response during the local accelerated phase of the pandemic Figure 1 Legend. (A) Northwestern Medicine COVID-19 in-patient census from 3/1/2020 through 8/6/2020. (B) Chicago COVID-19 case rate by date. Cases presented as case/100,000 population.

### Measures

The baseline survey collected self-reported data on: demographics (age, gender, race/ethnicity, job position, and home zip code); medical history and comorbidities; history of COVID symptoms; history of SARS-CoV-2 testing and diagnosis of COVID-19; healthcare and non-healthcare exposures to COVID-19; work-related tasks; whether respondent cared for COVID-19 patients; the use of PPE during exposures. Participants were categorized into four broad occupational classes: 1) physicians; 2) nurses; 3) administrators; and 4) other occupations (e.g. Supplemental Table 1). The survey was developed using adapted questions from the World Health Organization COVID-19 HCW and seroprevalence protocols(5).

### Statistical Analysis

We estimated the demographics of the sampling frame by creating a weighted average of demographic data provided by the NM Human Resources Department (NM employees) and McGaw Medical Center (NM residents and fellows). Due to under-representation of Hispanic and Non-Hispanic Black participants in our cohort, we used inverse probability weighting to estimate the prevalence of IgG positive serologic status within NM HCWs.

To create stable estimates in statistical models and preserve participant anonymity, several occupation groups with <50 participants were pooled together based on the research team’s perceived likelihood of SARS-CoV-2 exposure at work for each group (i.e., similarity in degree of patient contact and tasks performed; See Supplemental Table 1).

Up to 2.2% of survey participants had at least one incomplete question for demographics, occupation group, and patient care-related tasks and 4% were missing responses for symptoms. We excluded participants with missing data when the missing variable was the primary exposure of interest in a given model.

The prevalence and 95% confidence intervals (CIs) of HCWs with positive IgG antibodies to SARS-CoV-2 were calculated using exact binomial methods and described by age, gender, race/ethnicity, job position, patient care tasks, and COVID-19 exposures. Administrators were included as the referent group in occupation analyses to reflect exposure consistent with non-HCWs. To assess the independent associations between each of the covariates and seropositivity, we calculated odds ratios (ORs) and 95% CI from logistic regression models adjusted for age, sex, race/ethnicity, and self-reported out-of-hospital exposure to COVID-19. We used Holm’s procedures to adjust for multiple testing in the 14 patient care task groups and symptom questions.(6) The influence of variability in community spread was investigated by mapping employees’ residential address zip code with Illinois Department of Public Health COVID-19 case reporting data from June 15, 2020. All analyses were conducted using R software, version 3.6.0 (R Core Team, 2019).

## Results

The cohort included 79.6% women, and 74.9% non-Hispanic white, 9.7% Asian, 7.3% Hispanic, and 3.1% non-Hispanic Black participants; the mean (SD) age was 40.6 (12.0) years. The largest occupation groups sampled were nurses (n=1,794), physicians (n=1,260), and administrators (n=904). The demographics of occupation groups are shown in Table 1 and Supplemental Table 1.

The crude overall prevalence rate of anti-SARS CoV-2 IgG positive status was 4.8% (95% confidence interval [CI]: 4.6%-5.2%). The inverse probability weighted value (adjusted for the race/ethnicity of the sampling frame) was 5.3% (95% CI: 4.8%-5.9%).

### Sociodemographics by Seropositivity

Participants between 18- and 29-years-old had higher seropositive rates than older age groups (7.4%, [95% CI: 6.1%-9.0%] vs. 4.2% [95% CI: 3.7%-4.8%] (Table 2). Hispanic and non-Hispanic Black participants had the highest IgG+ prevalence rates of 9.6% (7.1%-12.7%) and 8.5% (5%-13.2%), respectively. Asian and White HCWs had prevalence rates of 4.6% (3.1%-6.5%), and 4.3% (3.8%-5.0%), respectively. There were no significant differences in the seropositive rates across gender or self-reported history of diabetes, hypertension, and obesity.

**Table 2:**
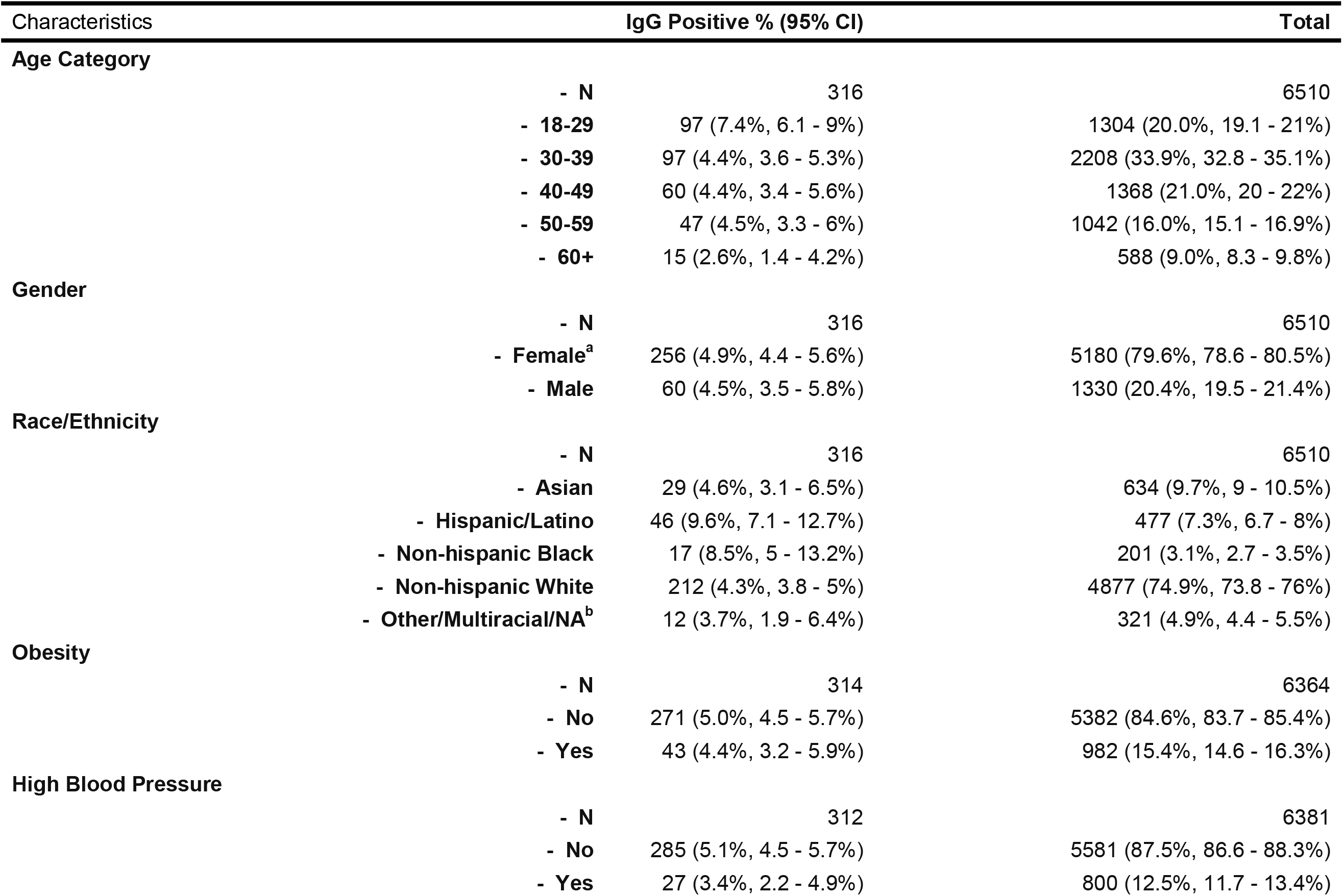
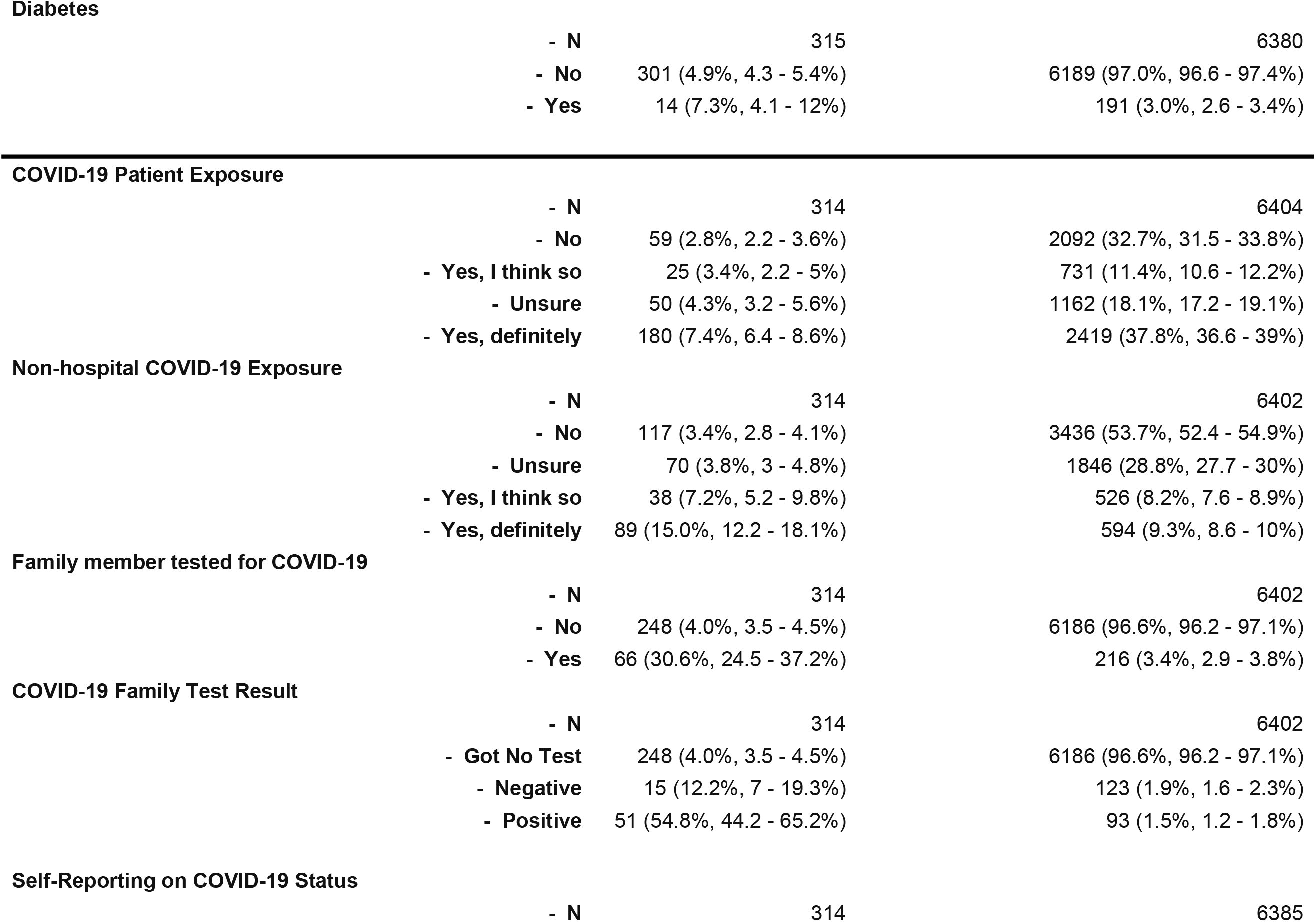
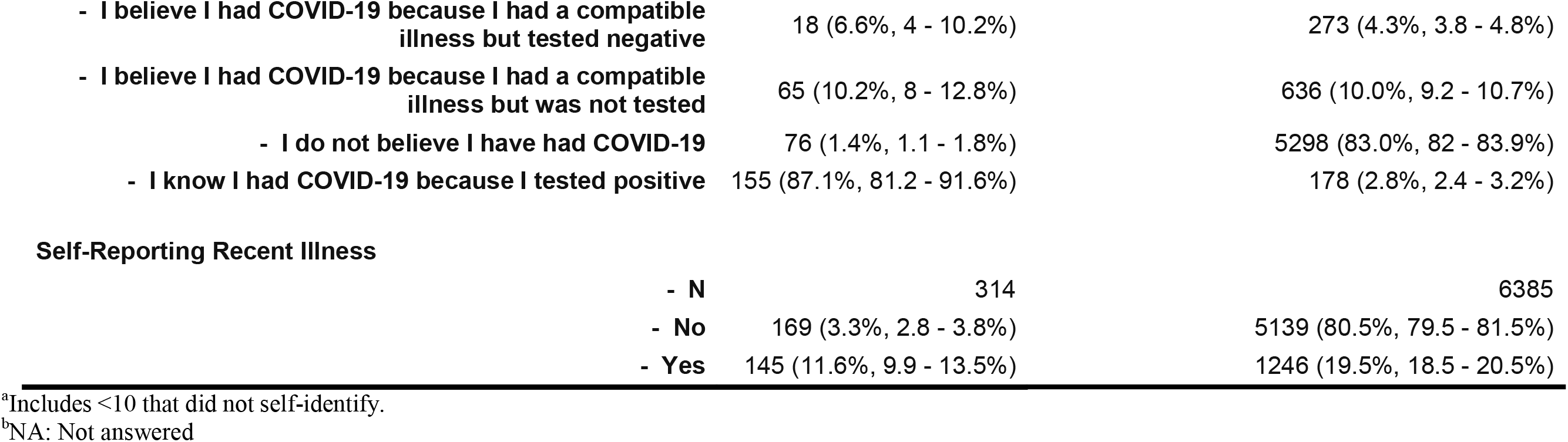
Participant Characteristics By Seropositive Group

### Out-of-hospital Exposure

Participants who reported a known out-of-hospital exposure (9.3%) had a seropositive rate of 15.0% (95% CI:12.2-18.1%). Those who reported having a family member in their home residence who tested positive for COVID-19 (n=93) had seropositive rates of 54.8% (44.2%-65.2%). After demographic adjustment, the adjusted OR for seropositive status of participants with a known out-of-hospital exposure was OR=4.7 (95% CI: 3.5-6.4) when compared with those without. Participants with a family member who tested positive for COVID-19 had demographic-adjusted OR=26.8 (17.3-41.8) when compared with those without a positive family member.

### Occupation Categories

Across occupation groups we observed crude prevalence rates of 10.4% (95% CI: 4.6-19.4%) in support service HCWs (i.e., environmental services, food services, and patient transporters) and 10.1% (5.5%-16.6%) in medical assistants. Nurses and respiratory technicians had crude seropositive rates of 7.6% (6.4%-9.0%) and 9.3% (3.1%-20.3%), respectively. Administrators had crude seropositive rates of 3.8% (2.6%-5.2%) and physicians had rates of 3.3% (2.3%–4.4%).

In unadjusted models, support services, medical assistants, and nurses had higher odds for being seropositive (as compared with administrators) of OR=3.0 (95% CI: 1.2-6.4), 2.9 (1.4-5.5), and 2.12 (1.5-3.2), respectively (Figure 2). After adjustment for demographics and self-reported out-of-hospital exposure to someone with COVID-19, the association remained significant for nurses (OR=1.9, 1.3-2.9), but was no longer significant for all other occupation groups.

**Figure 2:**
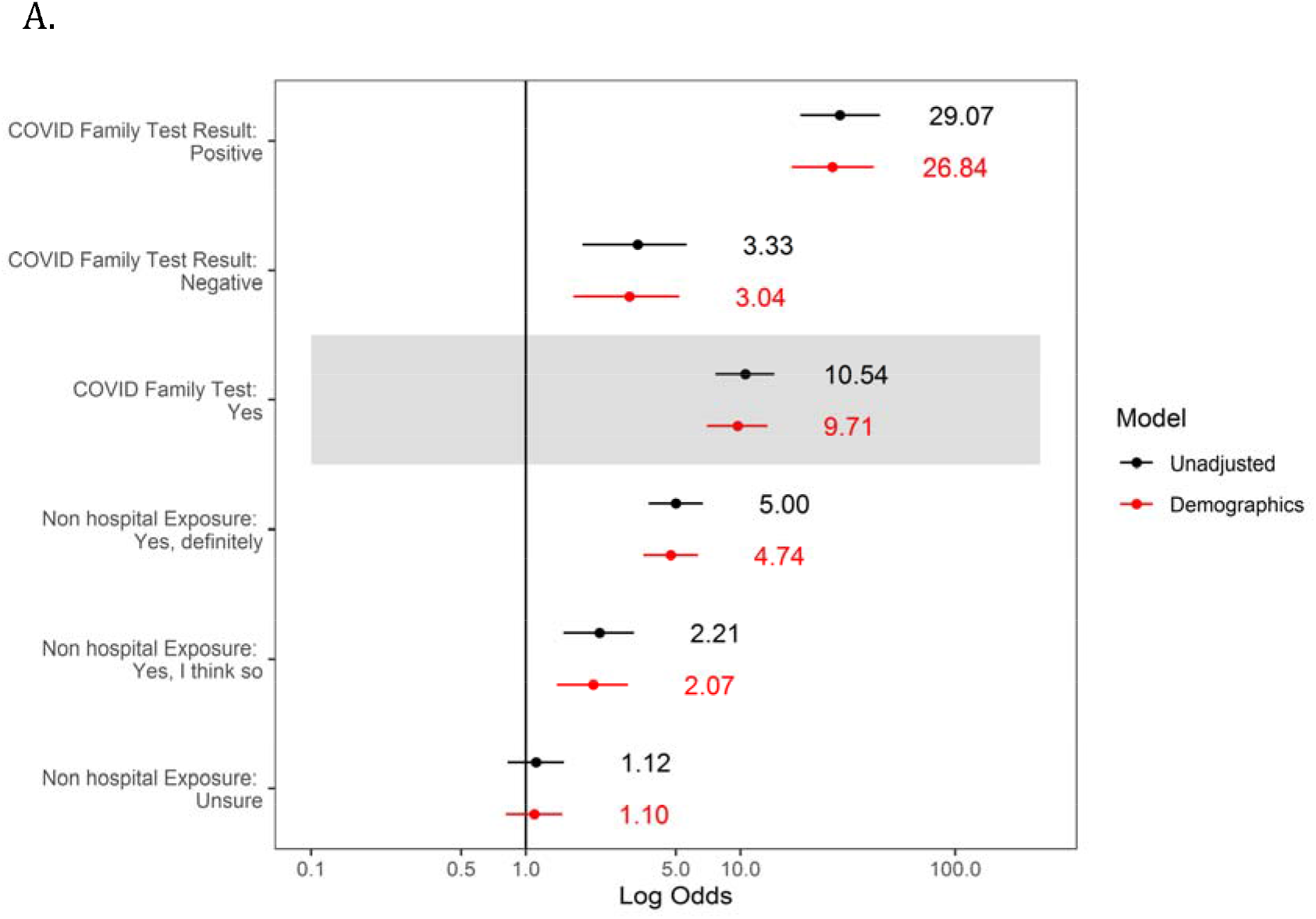
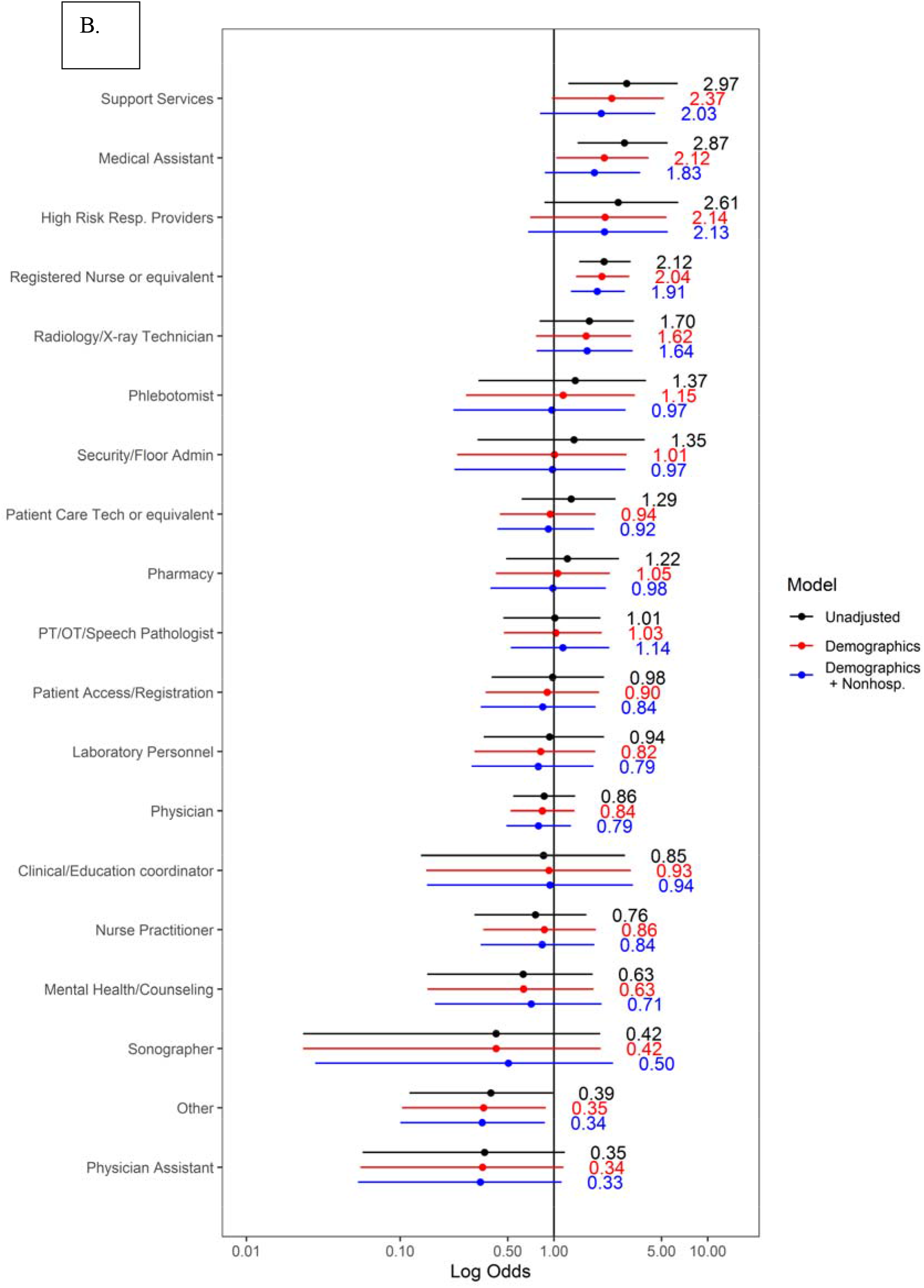
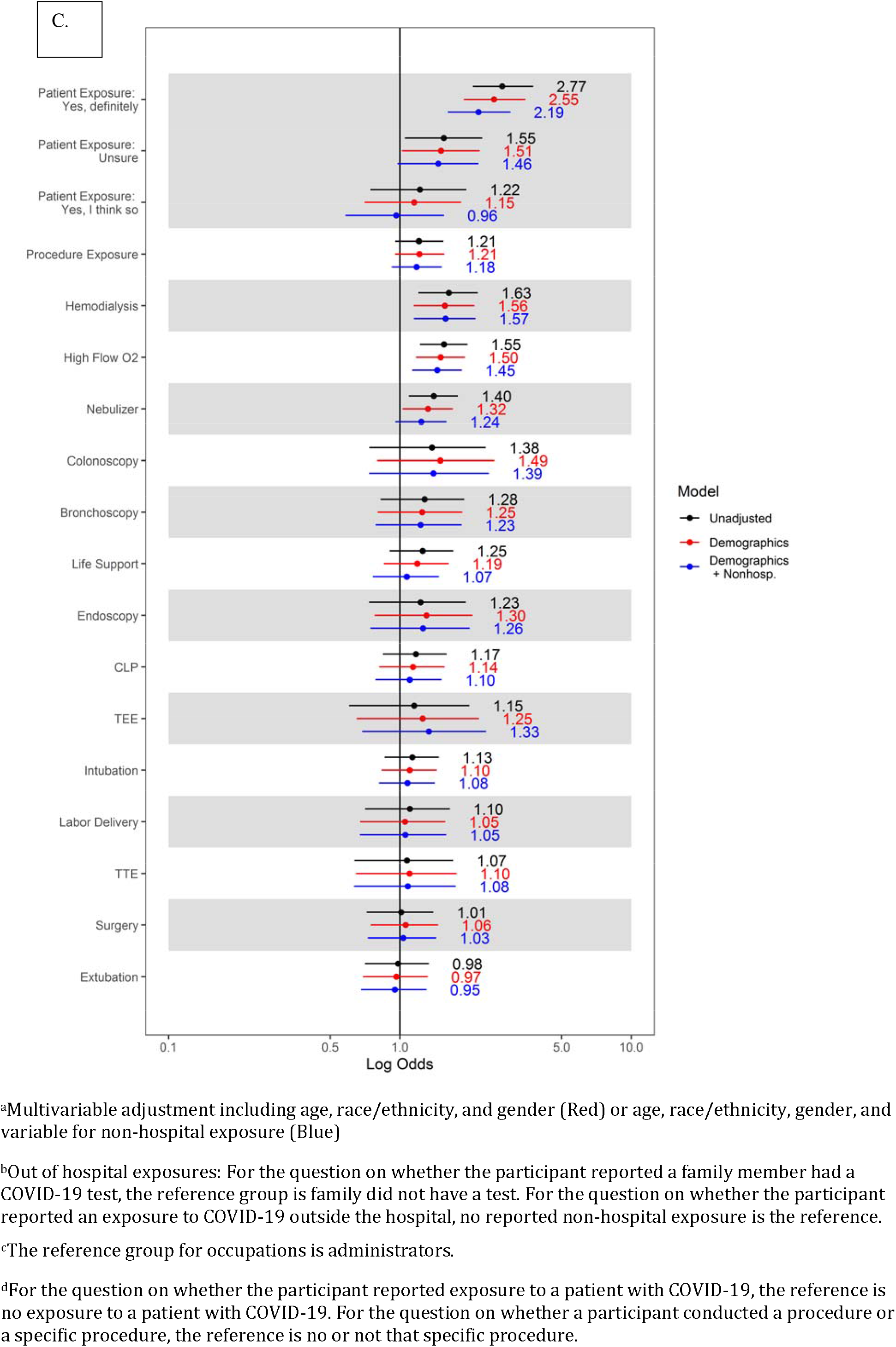
Unadjusted and multivariable adjusted^a^ logistic regression models of the association between anti-SARS-CoV-2 Seropositive Status and (A) Out-of-Hospital Exposures’^b^, (B) Occupation Group^c^, and (C) Clinical Care Tasks^d^.

Among physician specialties, the seropositive prevalence rate was 6.4% (95% CI: 3.1%-11.5%) for surgeons, 6.0% (1.7%-14.6%) for anesthesiologists, 4.3% (0.9%-12.2%) for Emergency Medicine, 2.9% (1.6%-5%) for Medicine and Family Medicine, and 0.5% (0-2.6%) for pediatrics. (Supplemental Table 2) Among Medicine subspecialties, the seropositivity rate for pulmonary/critical care (N=34) was 0% (0-10.3%).

### Occupational Tasks

Significantly higher crude rates for IgG+ were seen in HCWs who reported being exposed to COVID-19 patients (n=2,419) than those who did not (7.4%; 95% CI 6.4%-8.6% vs. 2.8%; 95% CI: 2.2%-3.6%). Among HCWs who were involved in overall patient care, those exposed to patients receiving high-flow oxygen (n=1,842) and nebulizer therapy (n=1,653) had higher rates of seropositive status (6.4% [5.3%-7.6%] vs. 4.2% [3.7%-4.9%]) and (6.1% [5.0%-7.4%] vs. 4.4% [3.9%-5.1%]), respectively, than those who were not. Exposure to patients receiving hemodialysis (n=807) was also associated with higher crude seropositive status rates (7.2% [5.5-9.2] vs. 4.5% [4.0-5.1%]). Intubation, bronchoscopy, and surgery were not significantly associated with seropositivity.

In demographic- and out-of-hospital-adjusted models, participating in the care of COVID-19 patients remained associated with higher seropositivity (OR=2.19 [95% CI:1.61-3.01]) when compared with those who did not report participating in the care of COVID-19 patients. Being exposed to patients receiving high-flow oxygen therapy, and hemodialysis also remained significantly associated with a 45% and 57% higher odds for seropositive status, respectively. Participation in transesophageal echocardiography (n=214), intubation (n=1360), and bronchoscopy (n=431) were not significantly associated with seropositive status when compared to participants who did not participate in those procedures.

### Community Variation in Seropositivity

The percent seropositive status by Chicago-neighborhood is shown in Figure 3. The highest case rates were in the Southwest and Northwest-side neighborhoods and lower case rates on the North Side and near-north suburbs. The neighborhood of residence of study participants and COVID-19 seropositivity mirrors COVID-19 case rates in the Chicago-land area.

**Figure 3:**
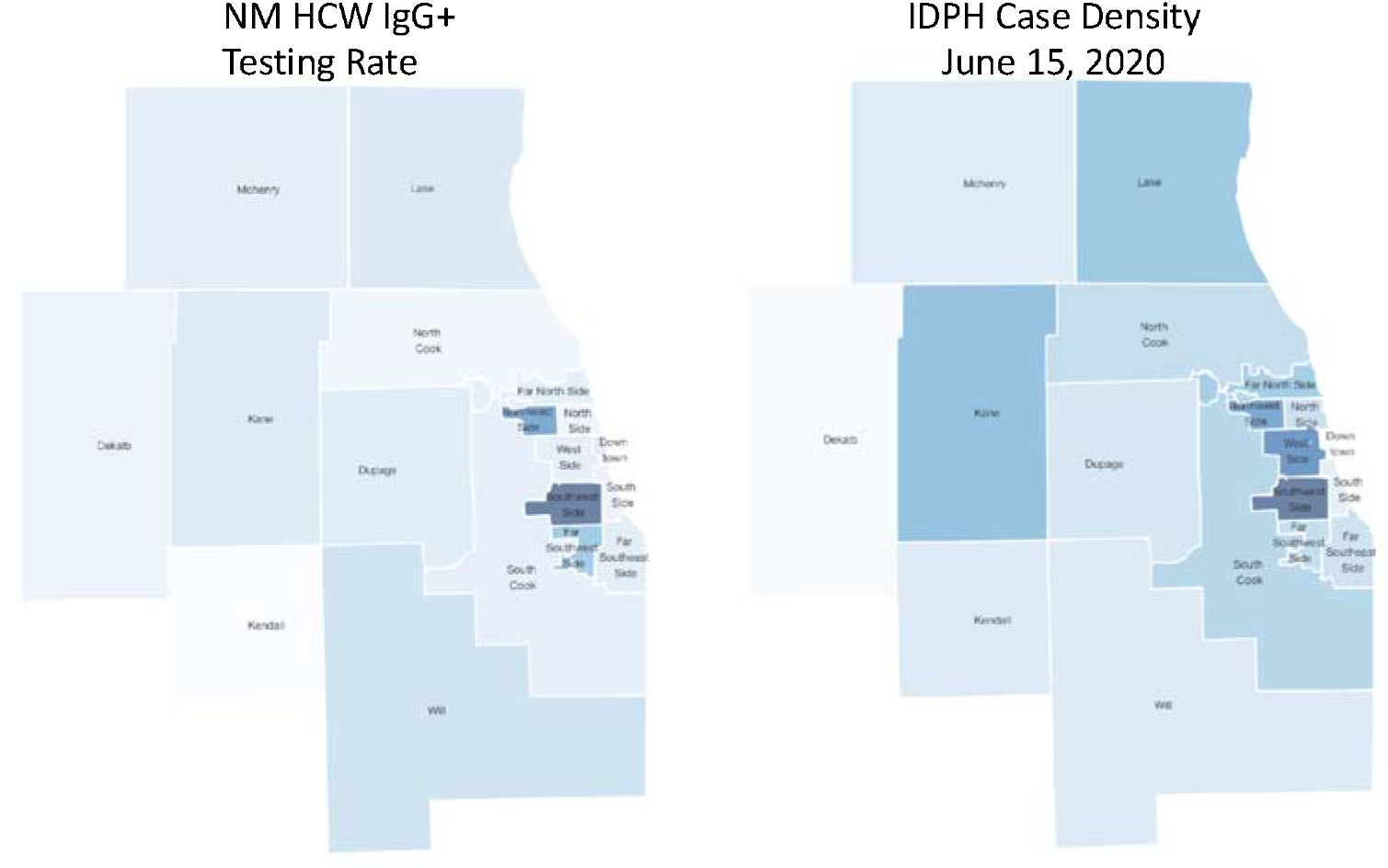
Chicago Neighborhoods and Surrounding Counties Heat Map by Seropositive Rate for NM HCW and Chicagoland COVID-19 Case-Rate Data Figure 3 Legend. Range of % positive IgG across neighborhoods on the left. COVID-19 case rates from the Illinois Department of Public Health as of June 15, 2020 (right). Darker colors represent higher IgG/case % rate, lighter represent lower IgG/case + rates.

### Reporting on Previous Infection and Impact on Health

Participants who reported that they did not believe that they had been infected with COVID-19 (n=5,298, 83%) had an IgG seropositive rate of 1.4% (95% CI: 1.1-1.8%), (n=76). These 76 participants represented 24% of all seropositive participants in the study. Participants who thought they might have been infected with COVID-19 but tested PCR negative, or were not tested for virus, had seropositive rates of 6.6% (4.0%-10.2%) and 10.2% (8.0%-12.8%), respectively. Participants who reported that they knew they had COVID-19 because they had a positive PCR test had a seroprevalence rate of 87.1% (81.2%-91.6%).

Among all seropositive participants in the study, 145 (46.2%) reported having a decline in their health. Seropositivity varied by symptoms with loss of smell or taste (OR=13.2, [9.8-17.8]) having the strongest association with positivity (Supplemental Figure2).

## Discussion

Within a single large health system serving Northeastern IL, we observed substantial variability in seropositivity rates by occupational class and tasks. However, despite these clear risks within the healthcare setting, out-of-hospital (community and home) exposure had the largest association with seropositive status.

Of all occupation groups, only nurses had higher odds ratios for seropositive status in demographic and out-of-hospital adjusted models. The higher risks observed in nurses are likely a function of nurses’ essential role on the care team that relies on frequent and close contact with patients.(7) Socialization between HCWs, particularly localized groups like nurses, is another plausible vehicle for transmission, which may lead to “clusters” of infected HCWs within specific occupation groups that co-locate for meals or face-to-face meetings.(8) Our sample was 80% female, and similar to US healthcare worker statistics.(9) Although no difference in gender was identified in seropositivity, it is important to note that HCWs, and especially nurses, are overwhelmingly women, thus the burden of SARS-CoV-2 infection will be mostly borne by female HCWs.

Exposure to patients receiving hemodialysis and high-flow oxygen therapy were strong predictors of seropositive status, that may, in part, be because they are both sustained exposures for HCWs. Thus, differences by occupation group in exposure risk in healthcare settings may be due to risk for aerosolization and the duration of exposure to a patient with COVID-19.(10) This suggests that availability and appropriate use of PPE and diligent infection control procedures can keep HCWs safe during brief exposures, while more work is needed on how to sustain protection over longer term exposures.

Approximately 1 in 5 participants who were seropositive did not think they had COVID-19, which is consistent with prior estimates of asymptomatic rates of COVID-19 infection that have ranged from 20%-40% in the general population and among HCWs.(11) Many factors associated with COVID-19 infection in community surveillance studies were correlated with HCW seropositive status. For example, we observed higher rates in Hispanic and non-Hispanic Black HCW cohort participants. In Chicago, COVID-19 case rates are higher, on average, in neighborhoods with a higher proportion of Black and Hispanic residents.(3,12) Detailed study of the socioeconomic characteristics, modifiable behaviors, and community events that facilitate virus transmission in these neighborhoods needs to be undertaken.

There are some important limitations to this study. First, these data represent a single, large health system that maintained adequate PPE throughout the crisis and launched infection control policies early on. Thus, the findings may not be generalizable to hospital systems working in communities where the burden exceeded the health system capacity. Second, while the seroprevalence reporting by race and ethnicity is consistent with national reports describing higher rates of infection in Black and Hispanic adults, we had relatively small numbers of these groups in our sample and so estimates may be unstable. Third, our data on occupation group and work-related behaviors come from survey data, which may be susceptible to recall bias, particularly in participants who received their serologic testing results prior to filling out their surveys. We did not, however, see different directions of association between work task, location, and risk for prevalent COVID-19 when we stratified the cohort by the relative timing of serologic testing and questionnaire completion, suggesting that recall bias does not explain the reported associations between work type, symptoms, and beliefs about COVID-19 infection and serologic status. Fourth, the performance of all currently-available assays for IgG detection have not been rigorously validated in community-based studies with consistent reference standard samples. Further, some individuals infected with SARS-CoV-2 may not develop a detectable antibody response, and/or their serum antibody presence may be transient.(13) Thus, the reported prevalence estimates could be under- or over-estimated if the accuracy and precision of the assays were lower than initially reported. However, the relative differences that we observed across groups would not be systematically biased by assay performance alone.

In conclusion, HCWs in this study were at modestly lower risks for SARS-CoV-2 infection as compared with other studies of HCWs from the New York area and Spain, and similar seropositive rates as reported in Denmark.(14-16) Across occupation groups, nurses were at the highest-level risk from work-related exposures. Given the exposure that HCWs face in the direct care of patients with known and unknown COVID-19 status, these data support the effectiveness of PPE and infection control policies to keep HCWs safe. In the setting of a well-resourced health system not overwhelmed by hospitalized COVID-19 patients, the majority of risk for SARS-CoV-2 infection was associated with community transmission, suggesting that persistent infection control within the workplace will require adequate PPE supplies, refined infection control policies, and sustained vigilance in and out of the hospital.

## Data Availability

The data are not available for public use.

## Acknowledgements

We would like to thank our colleagues at Northwestern Medicine who helped coordinate the serologic testing effort for NM employees and data extraction. In particular we would like to thank Jay Anderson, Julia Lynch, Anne Cunningham, F. Andy Eichler, Kenneth Hedley, Kristina Hedley, Jen Steinmetz, and Tracey Woods. Thank you to informatics colleagues Quan Mai, Daniel Schneider, Theresa Walunas, Firas Wehbe. We would like to thank the Illinois Department of Public Health for providing files of the public data, the Northwestern All of Us Research Program for providing staff, and Dr. Donald M. Lloyd-Jones for his editing of this manuscript. We are grateful to the NM employees who volunteered to participate in this project.

---

**Supplemental Table 1:**
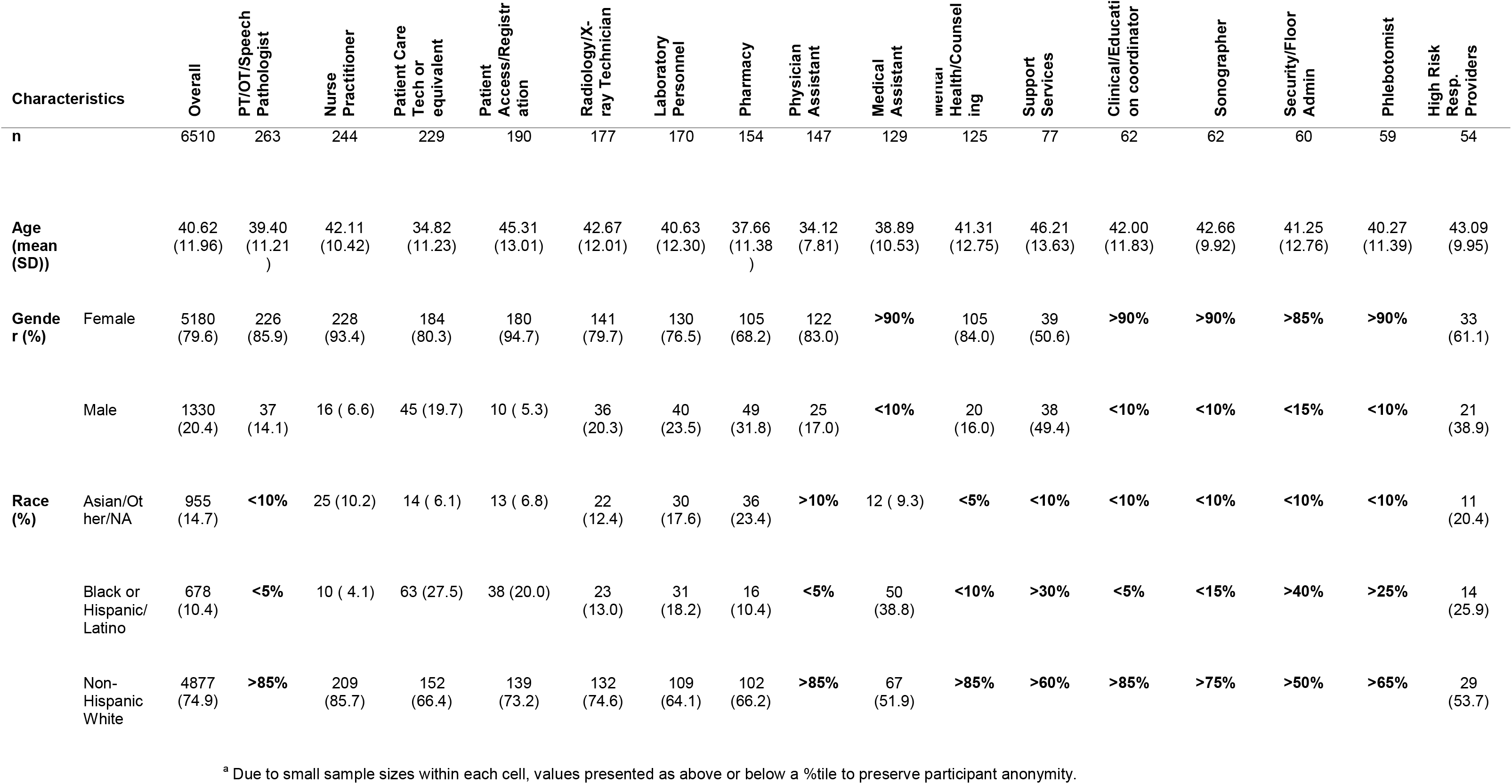
Demographic Characteristics of Occupation Groups of less than 500 participants.^a^

|  |  |  |  |  |  |  |  |  |  |  |  |  |  |  |  |  |  |  |
| --- | --- | --- | --- | --- | --- | --- | --- | --- | --- | --- | --- | --- | --- | --- | --- | --- | --- | --- |
| Characteristics |  | Overall | PT/OT/Speech Pathologist | Nurse Practitioner | Patient Care Tech or equivalent | Patient Access/Registration | Radiology/X-ray Technician | Laboratory Personnel | Pharmacy | Physician Assistant | Medical Assistant | Mental Health/Counseling | Support Services | Clinical/Educational coordinator | Sonographer | Security/Floor Admin | Phlebotomist | High Risk Resp. Providers |
| n |  | 6510 | 263 | 244 | 229 | 190 | 177 | 170 | 154 | 147 | 129 | 125 | 77 | 62 | 62 | 60 | 59 | 54 |
| Age (mean (SD)) |  | 40.62 (11.96) | 39.40 (11.21) | 42.11 (10.42) | 34.82 (11.23) | 45.31 (13.01) | 42.67 (12.01) | 40.63 (12.30) | 37.66 (11.38) | 34.12 (7.81) | 38.89 (10.53) | 41.31 (12.75) | 46.21 (13.63) | 42.00 (11.83) | 42.66 (9.92) | 41.25 (12.76) | 40.27 (11.39) | 43.09 (9.95) |
| Gender (%) | Female | 5180 (79.6) | 226 (85.9) | 228 (93.4) | 184 (80.3) | 180 (94.7) | 141 (79.7) | 130 (76.5) | 105 (68.2) | 122 (83.0) | >90% | 105 (84.0) | 39 (50.6) | >90% | >90% | >85% | >90% | 33 (61.1) |
|  | Male | 1330 (20.4) | 37 (14.1) | 16 ( 6.6) | 45 (19.7) | 10 ( 5.3) | 36 (20.3) | 40 (23.5) | 49 (31.8) | 25 (17.0) | <10% | 20 (16.0) | 38 (49.4) | <10% | <10% | <15% | <10% | 21 (38.9) |
| Race (%) | Asian/Other/NA | 955 (14.7) | <10% | 25 (10.2) | 14 ( 6.1) | 13 ( 6.8) | 22 (12.4) | 30 (17.6) | 36 (23.4) | >10% | 12 ( 9.3) | <5% | <10% | <10% | <10% | <10% | <10% | 11 (20.4) |
|  | Black or Hispanic/Latino | 678 (10.4) | <5% | 10 ( 4.1) | 63 (27.5) | 38 (20.0) | 23 (13.0) | 31 (18.2) | 16 (10.4) | <5% | 50 (38.8) | <10% | >30% | <5% | <15% | >40% | >25% | 14 (25.9) |
|  | Non-Hispanic White | 4877 (74.9) | >85% | 209 (85.7) | 152 (66.4) | 139 (73.2) | 132 (74.6) | 109 (64.1) | 102 (66.2) | >85% | 67 (51.9) | >85% | >60% | >85% | >75% | >50% | >65% | 29 (53.7) |

<sup>a</sup> Due to small sample sizes within each cell, values presented as above or below a %tile to preserve participant anonymity.

**Supplemental Table 2:**
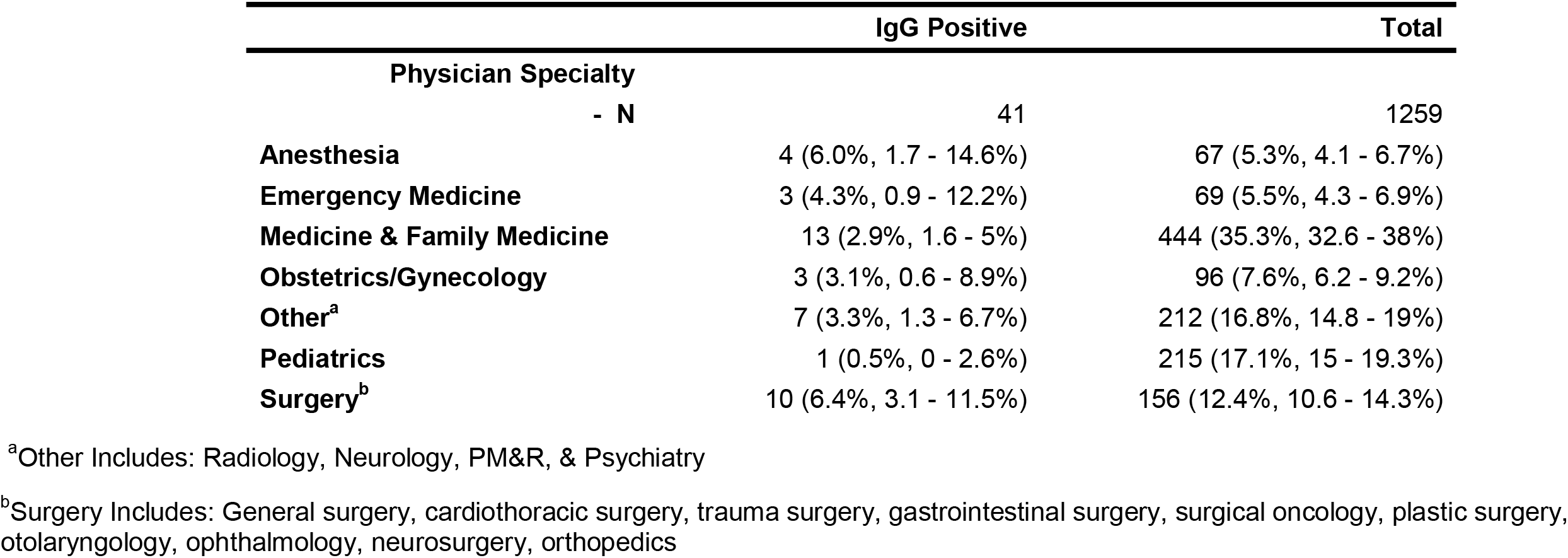
Crude Seropositive Rates By Medical Specialty

**Supplemental Figure 1:**
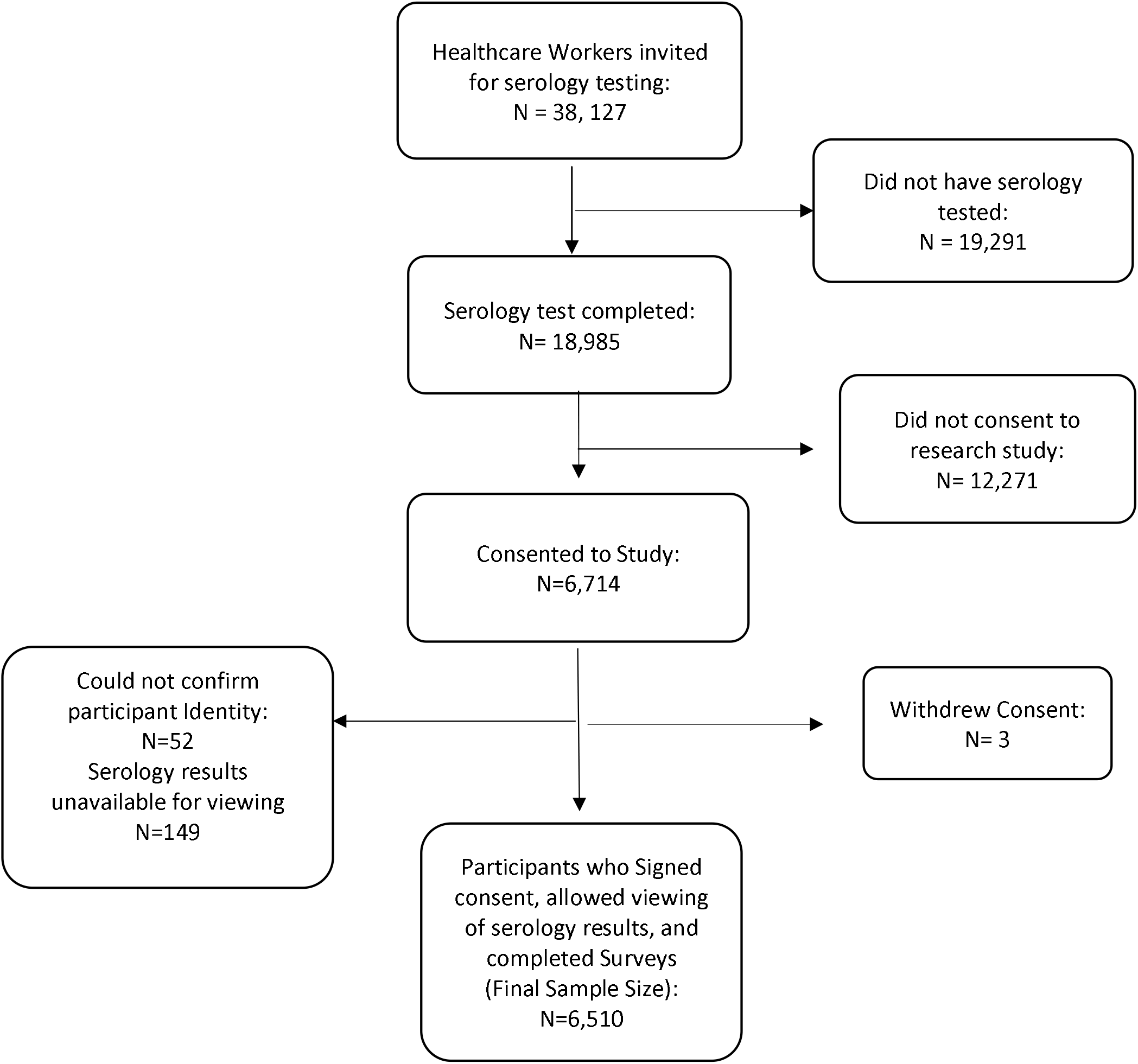
Enrollment Flowchart

**Supplemental Figure 2:**
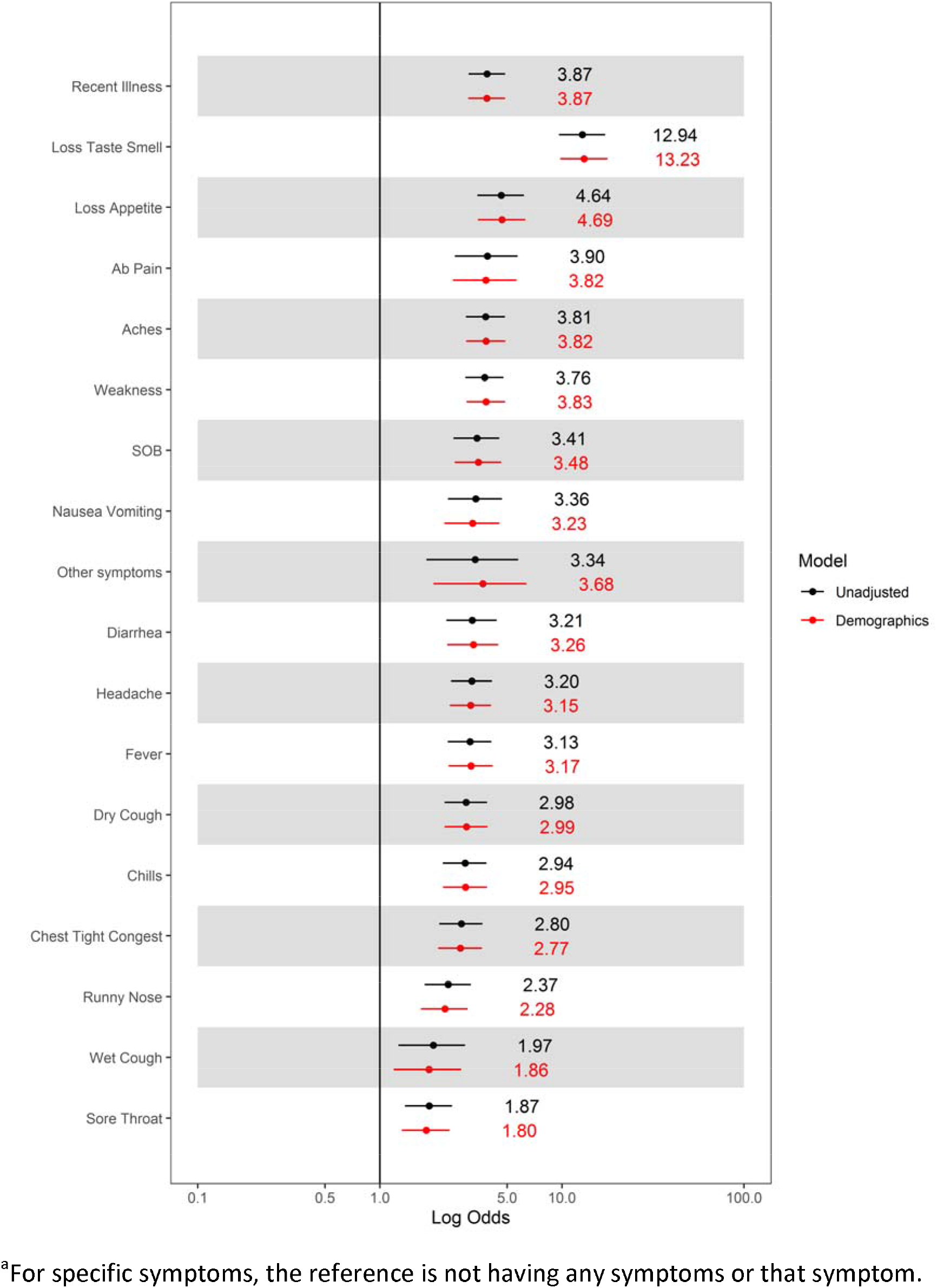
Unadjusted Logistic Regression Models for Symptoms and SARS CoV-2 IgG+ Serologic Status among Northwestern Medicine Healthcare Workers^a^

